# The Use/Abuse of Oral Contraceptive Pills Among Males: A Mixed-Method Explanatory Sequential Study Over Jordanian Community Pharmacists

**DOI:** 10.1101/2021.03.11.21253403

**Authors:** Muna M. Barakat, Raja’a A. Al-Qudah, Amal Akour, Mona Abu-Asal, Samar Thiab, Yahya H. Dallal Bashi

## Abstract

**Background:** Oral contraceptive pills (OCPs) are considered one of the most important birth control methods in the world. However, these pills were designed for female administration rather than males. This study was designed to investigate patterns of OCPs use and abuse among Jordanian males, according to the community pharmacists’ observations.

**Method:** A mixed-method explanatory sequential study was conducted using an online self - administered survey and semi-structured in-depth interviews for community pharmacists. The interviews were utilized using a conceptual framework. Inductive thematic analysis and descriptive/regression analyses were completed using Nvivo and SPSS, respectively.

**Results:** A total of 158 questionnaire responses and 22 interviews were included in our analysis. Around half (48.4%) of the questionnaire responses confirmed that males could use OCPs for hair growth enhancement, muscle gain and acne treatment 12.7%, 31.7% and 4.4%, respectively. Through the interviews, the majority of the pharmacists highlighted that most of the males use OCPs for bodybuilding purposes, according to recommendations by their coaches at the gym. The most abused OCPs containing estrogen (Ethinyl estradiol) and progestins (Drospirenone or Levonorgestrel).

**Conclusion:** This study provided insight into unexpected uses of OCPs by males in Jordan. Community pharmacists have a crucial role in the management of OCPs use and abuse, However, restricted regulations and monitoring must be released and implemented on the community to limit such practices.

## 1. Introduction

The non-therapeutic consumption of medications, in terms of abuse and misuse, is recently on the rise. Medication abuse is defined as the usage of drugs for non-medical purposes [1]. The most common medications which are prone to abuse are products containing opioids, stimulants and laxatives [2-4]. However, the abuse of oral contraceptive pills (OCPs) has not been fully examined yet.

OCPs are one of the most common birth control methods in the United States (US), mainly used by females between the ages of 25-44 years old [5]. There are three types of OCPs: combined estrogen-progesterone, progesterone alone and continuous dosing or extended cycle pills [6]. It was reported that the combined oral contraceptive (COC) pills were the most prescribed compared with the other types [6]. The main indication for these pills is pregnancy control; however, the Food and Drug Association (FDA) labeled additional uses for them such as menstrual period disorders (Dysmenorrhea, amenorrhea, oligomenorrhea), acne treatment, hirsutism, polycystic ovary syndrome and many other uses [7]. OCP’s labeled indications are generally approved for females rather than males. The frequent use of OCPs among males may contribute to adverse physiological effects such as gynecomastia, testicle shrinkage, prostate cancer and diminished libido [8].

In Jordan, it was reported that OCPs are the second most common birth control method [9, 10], and OCPs are sold over the counter [9]. Such unsupervised practice could be associated with inappropriate administration of many medications [11] including OCPs, as reported by the Jordanian community pharmacist in a previous study [12]. In that study, pharmacists stated that both genders are prone to OCPs improper usage, such as topical application of OCPs for hair growth enhancement and OCPs were used to give a false negative result for an addictive drug screening test [12]. Therefore, this study aims to investigate the patterns of OCPs use and abuse among Jordanian males, according to the community pharmacists’ observations.

## 2. Method

A mixed-method explanatory sequential (qualitative and quantitative) study was conducted in order to understand the pharmacist observations about patterns of OCPs use/abuse among Jordanian males. The first phase of the study, which was conducted from March 15^th^ to April 24^th,^ 2020, consisted of a self-administered online survey, targeting Jordanian community pharmacists and pharmacy trainees. Pharmacists were recruited through social media platforms (Facebook, WhatsApp, LinkedIn, and Twitter). The second phase of the study, which was carried out from April 25^th^ to May 15^th^, 2020, consisted of online semi-structured in-depth interviews, using Zoom®, with maximum variation purposive sample of community pharmacists. The study was approved by the Institutional Review Board of Applied Science Private University [Ethical approval number: 2019-PHA-14]. The sequential method is represented in Figure 1.

**Figure 1.**
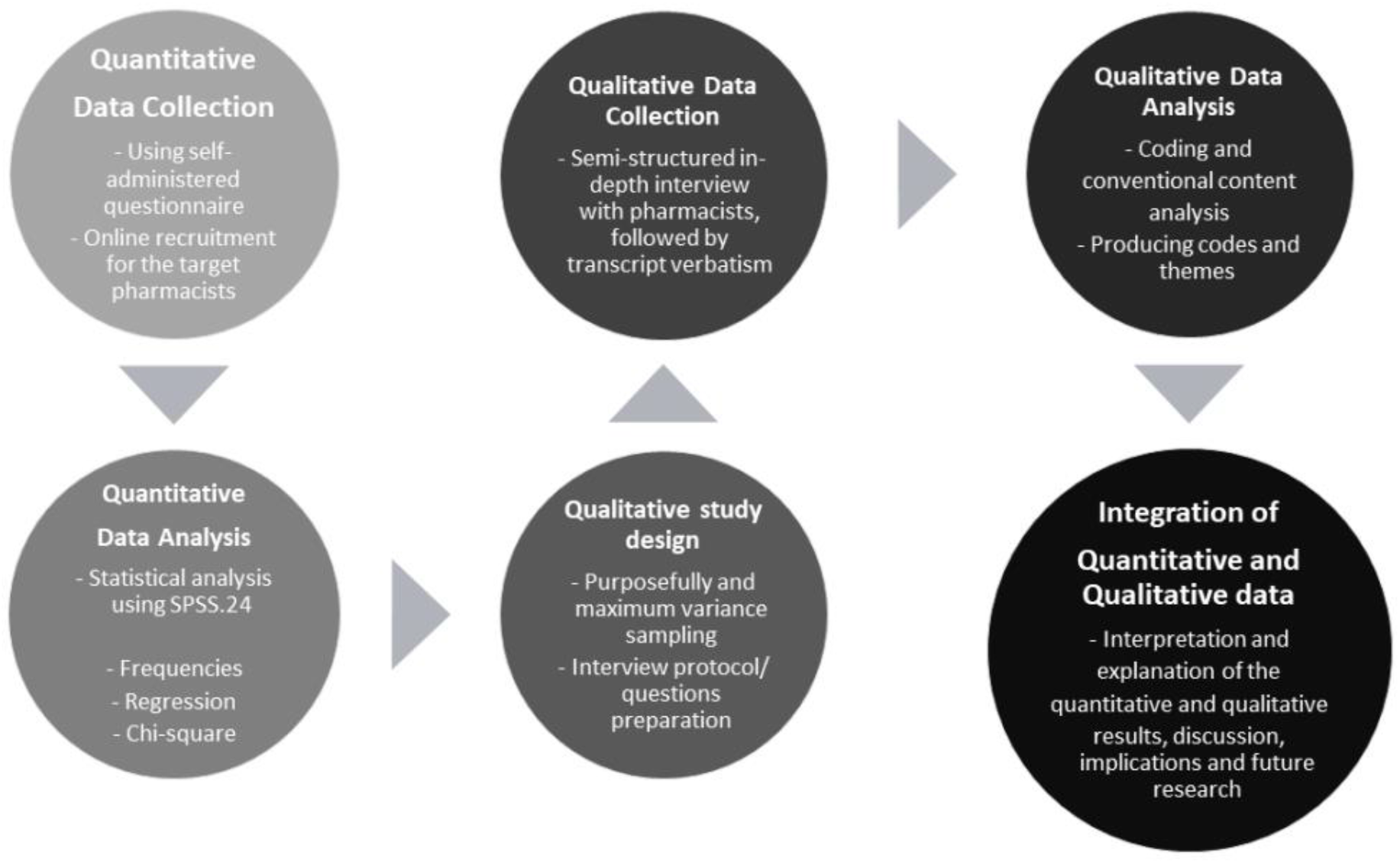
Descriptive scheme for the study mixed sequential method.

### 2.1 Quantitative method (first phase)

An online self-administered survey has been developed and validated (face and content validity) by clinical researchers to solicit anonymous responses, which were treated confidentially. The survey has been developed using the general principles of good survey design [13]. The surveys contain closed-ended and open-ended questions that are completed within an average of 7 minutes. The survey had been administered in Arabic and English via Google® forms technology. Participants were also clarified that their participation in the study was voluntary and did not pose any risks. Potential participants who completed the survey were considered to have given informed consent for the study participation.

The first draft of the survey was appraised by 20 independent academic staff members who have decent experience in oral contraceptives research studies; as well a statistician was consulted at this stage of validation. The final version of the survey was refined according to the provided comments and feedback, translated from English into Arabic and then was back-translated by two specialized academics. The questions were free from medical jargon or difficult terminologies. The final validation stage was the piloting step, which involved 25 academic and 25 non-academic participants. This stage of the study was conducted to enhance clarity, readability, understandability, and confirm the study applicability in the Jordanian community pharmacists. Internal consistency reliability was tested by the Cronbach’s alpha coefficient (=0.82).

The study survey included three parts. The first part (Part A) comprised nine questions including sociodemographic information; the second part (Part B) consisted of four questions comprising details of knowledge and beliefs of pharmacists about OCPs uses for males. The questions of knowledge were mainly concerned about the presence of possible indications for OCPs in males and the risks of using those pills. The last part (Part C) was focusing on the pharmacist experience and practice towards OCPs usage by males; which consisted of ten multiple-choice questions.

Based on Tabachnick and Fidell recommendation for sample size calculation in linear regression analysis, 5-20 subjects per predictor are suggested to be preferable [14]. Based on the number of independent variables levels used in this study (n = 9) and using the number of 10 subjects per predictor level, a minimum sample size of 90 or higher was considered suitable for the purpose of this study.

In this study, the recruited sample was 162 community pharmacists who met the inclusion criteria. The eligibility criteria for participation are being Jordanian pharmacists and trainees (who were working/training in community pharmacy). Sample-sized recruitment was conducted using a convenience sampling method. The inclusion criteria were explained at the start of the survey, which stated: “If you are working or training at a community pharmacy, please let us know if you would like to participate in this survey”.

### 2.2 Statistical analyses

The data of the completed surveys were extracted from an electronic platform and exported to Statistical Package for Social Sciences version 24.0 (SPSS® Inc., Chicago, IL, USA) for the statistical analysis. Descriptive statistics included percentages, means, and frequency distribution, calculated for each question. Descriptive and univariate correlation analyses using Pearson correlation coefficient (r) were used for the correlation, which was conducted at a 5% significance level. A *p*-value of < 0.05 represented a significant difference. Factors affecting the OCPs’ improper uses were analyzed using simple and multivariate linear regression.

### 2.3 Qualitative Method (Second Phase)

The qualitative study was conducted using semi-structured in-depth interviews for community pharmacists. The sample was recruited purposively then maximum variation sampling was conducted, to reach sample saturation (22 community pharmacists). Purposeful sampling directed the researcher to find community pharmacists with pure experiences regarding OCPs usage among males. In this regard, introductory pilot interviews were employed to find the pharmacists who provide fruitful and detailed responses then the selected pharmacists participated in in-depth interviews.

A series of questions regarding the use of OCPs among males were asked and answered freely by the participants. This research instrument was reviewed by a group of leading clinical pharmacy researchers to ensure the suitability of the measurement tool. The interview included three probing questions. The first was “From your experience, please describe how could males use the OCPs”, and the second was “What is the best way to limit the abuse of OCPs among males?”, each interview lasted for 25 minutes. The participants were informed that all the data will be treated confidentially and signed electronic informed consents. The interviews were recorded and transcribed verbatim. Transcripts were written in Arabic (the native language of Jordan), translated into English, checked for clarity and accuracy, uploaded to NVivo12 (qualitative data analysis software; QSR International Pty Ltd. Version 12) for coding and then analyzed using inductive thematic analysis using conceptual framework methods [15]. The inter-rater reliability was assessed using Cohen’s kappa. The kappa value for the three coders was 0.85, indicating good agreement between the coders. The different researchers independently re-checked the trustworthiness and the accuracy of transcription, translation and inter-coder reliability.

## 3. Results

### 3.1 Sociodemographic characteristics of the participants in both quantitative and qualitative phases

Out of the total 162 completed questionnaires (response rate =98%), four forms (2.4%) were excluded from the study due to incomplete responses, accordingly, 158 (97.5%) questionnaires were included in the analysis. The majority of the participating pharmacists were females (n=118, 74.2%) and around half of them (n=79, 49.7%) were <40 years old and holding both university education degrees (bachelor’s and postgraduates) (n=111, 70.2%).

About half of the pharmacists (n=75, 47.5%) have been working in community pharmacies for more than 10 years, which is located in the capital of Jordan “Amman” (n= 115, 72.3%). Around 74% (n=117) of the pharmacy’s customers were classified as middle social class, from the pharmacists’ perception. It is worth noting that 44.7% (n=71) of the pharmacies were adjacent to sports gyms (Table 1).

**Table 1.** Sociodemographic characteristics of the participants in both Quantitative (n=158) and Qualitative phases (n=22).

| Characteristic | Questionnaire Sample<br>n (%) | Interviews Sample<br>n (%) |
| --- | --- | --- |
| **Gender** |  |  |
| • Female | 118 (74.2) | 12 (54.5) |
| • Male | 41 (25.8) | 10 (45.5) |
| **Age (years)** |  |  |
| • 20-25 | 16 (10.1) | 5 (22.7) |
| • 26-40 | 79 (49.7) | 5 (22.7) |
| • 41-55 | 59 (37.1) | 7 (31.8) |
| • >55 | 5 (3.1) | 5 (22.7) |
| **Education** |  |  |
| • Bachelor's degree | 100 (63.3) | 17 (77.3) |
| • Pharmacy student +trained in a pharmacy | 35 (22.2) | 4 (18.2) |
| • Postgraduate degree | 11 (6.9) | 1 (4.5) |
| • Diploma | 12 (7.5) | 0 |
| **Years of experience** |  |  |
| • <5 | 35 (22.2) | 4 (18.2) |
| • 5 to 10 | 48 (30.4) | 4 (18.2) |
| • 11 to 15 | 58 (36.7) | 5 (22.7) |
| • 16-20 | 8 (5.0) | 5 (22.7) |
| • >20 | 9 (5.7) | 4 (18.2) |
| **Position in the pharmacy** |  |  |
| • Employee pharmacist | 94 (59.5) | 9 (40.9) |
| • Trainee in a pharmacy | 37 (23.4) | 6 (27.3) |
| • Pharmacy owner | 27 (17.6) | 7 (31.8) |
| **Province where you work** |  |  |
| • The capital of Jordan (Amman) | 115 (72.3) | 13 (59.1) |
| • East of Jordan | 9 (5.7) | 2 (9.1) |
| • West of Jordan | 6 (3.8) | 3 (13.6) |
| • North of Jordan | 16 (10.1) | 2 (9.1) |
| • South of Jordan | 1 (0.6) | 2 (9.1) |
| **The most common social class distribution of the pharmacy customers** |  |  |
| • Low | 21 (13.3) | 5 (22.7) |
| • Middle | 117 (74.0) | 13 (59.1) |
| • High | 20 (12.7) | 4 (18.2) |
| **The presence of sport gym around the pharmacy** |  |  |
| • Yes | 71 (44.7) | 14 (63.6) |
| • No | 63 (39.6) | 8 (36.4) |
| • Not sure | 24 (15.1) | 0 |

The sociodemographic data of the qualitative phase was representing the maximum variation sample. A total of 22 pharmacists were involved in the study. Females percentage was slightly predominant over males (n=12, 54.5%), less than one third were 41-55 years old (n=7, 31.8%) and (n=17, 77.3%) were holding a bachelor’s degree (Table 1).

### 3.2 Knowledge and beliefs of pharmacists about male’s usage of OCPs

The majority of the participants(n=77, 48.4%) agreed that OCPs could be used by males. The pharmacists reported that males could use OCPs for muscle gain purposes, enhancement of hair growth and acne treatment in 31.7%, 12.7% and 4.4%, respectively. Pharmacists’ knowledge about possible OCPs side effects, when used by males, was assessed. The most reported side effects were gynecomastia, decreased libido, mood changes, and increased body weight (69.8%, 64.8% 64.8%, 57.9%) respectively (Table 2).

**Table 2.**
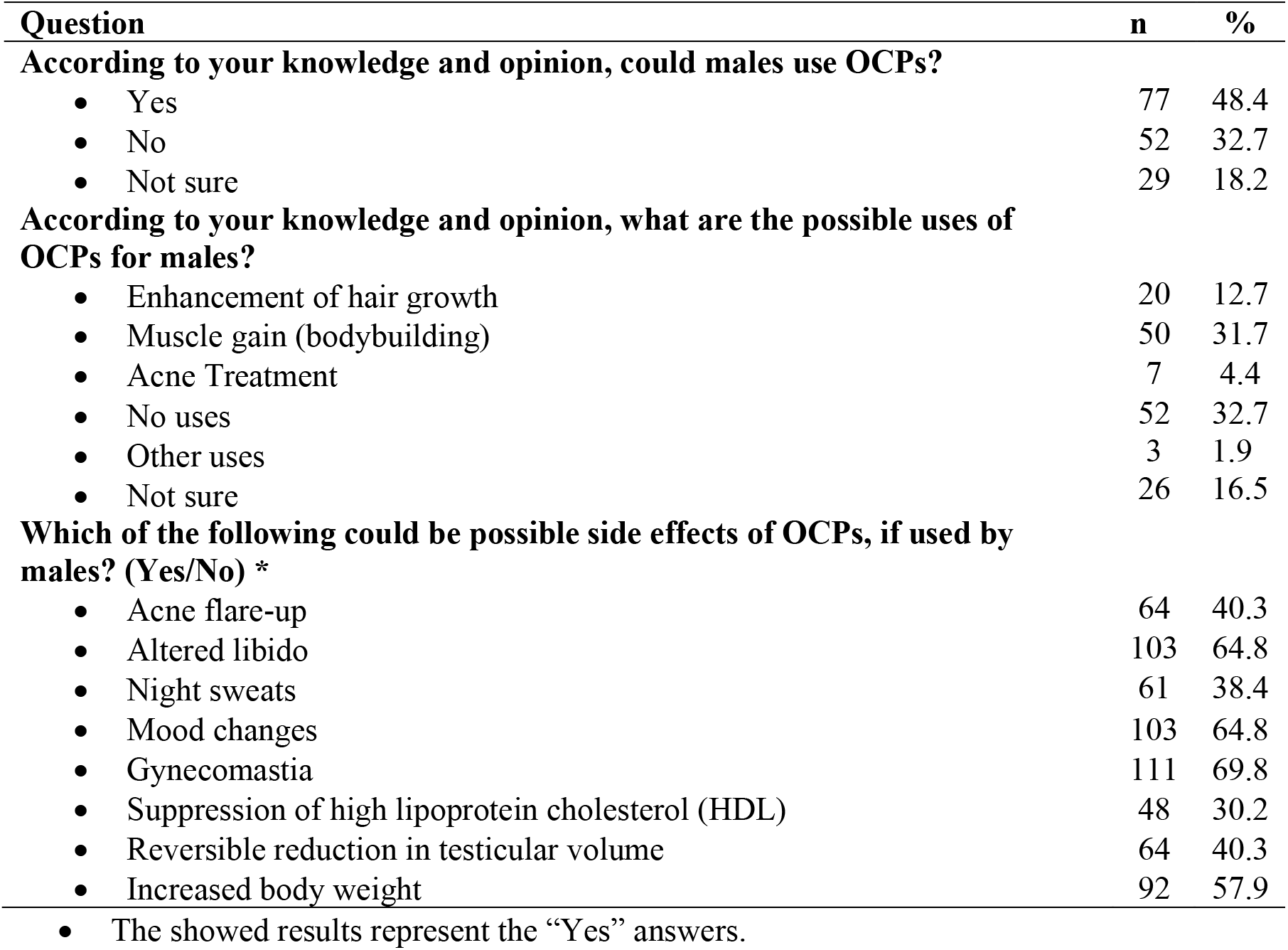
Pharmacists knowledge and beliefs about the possible uses of OCPs among males (n=158)

| **Question** | **n** | **%** |
| --- | --- | --- |
| **According to your knowledge and opinion, could males use OCPs?** |  |  |
| • Yes | 77 | 48.4 |
| • No | 52 | 32.7 |
| • Not sure | 29 | 18.2 |
| **According to your knowledge and opinion, what are the possible uses of OCPs for males?** |  |  |
| • Enhancement of hair growth | 20 | 12.7 |
| • Muscle gain (bodybuilding) | 50 | 31.7 |
| • Acne Treatment | 7 | 4.4 |
| • No uses | 52 | 32.7 |
| • Other uses | 3 | 1.9 |
| • Not sure | 26 | 16.5 |
| **Which of the following could be possible side effects of OCPs, if used by males? (Yes/No) \*** |  |  |
| • Acne flare-up | 64 | 40.3 |
| • Altered libido | 103 | 64.8 |
| • Night sweats | 61 | 38.4 |
| • Mood changes | 103 | 64.8 |
| • Gynecomastia | 111 | 69.8 |
| • Suppression of high lipoprotein cholesterol (HDL) | 48 | 30.2 |
| • Reversible reduction in testicular volume | 64 | 40.3 |
| • Increased body weight | 92 | 57.9 |

- The showed results represent the “Yes” answers.

### 3.3 Pharmacist experience and practice towards OCPs usage by males

Sixty-two percent of the participating pharmacists acknowledged that they dispense OCPs without prescription, and around 58% of them were dispensing OCPs for males for their personal use. In particular, for muscle gain (36.5%) and hair growth enhancement (20.8%). Some of the participating pharmacists also indicated that most males who used to use OCPs were stranger customers (n=43, 27.7%) and most commonly aged 20-35 (n=114, 71.7%). Study participants stated that the males’ OCPs users either directly ask and admit their need for these pills or are recognized by their facial expression and body language, 21.5% and 27.2%, respectively (Table 3).

**Table 3.** pharmacist experience and practice towards OCPs usage by males (n=158)

| **Question** | **n** | **%** |
| --- | --- | --- |
| **Have you ever dispensed OCP s in your pharmacy to males (for their personal use)** |  |  |
| • Yes | 92 | 57.9 |
| • No | 34 | 21.4 |
| • I have never asked about the user | 32 | 20.8 |
| **Have you ever dispensed OCPs to males for the following uses?** |  |  |
| • Enhancement for the hair growth | 32 | 20.8 |
| • Muscle gain (bodybuilding) | 58 | 36.5 |
| • Acne treatment | 2 | 1.3 |
| • Never | 66 | 41.5 |
| **The most common ages of males came to the pharmacy asking for OCPs (years)** |  |  |
| • < 20 | 12 | 8.2 |
| • 20-35 | 114 | 71.7 |
| • 36-50 | 30 | 18.9 |
| • > 50 | 2 | 1.3 |
| **Have you ever noticed any inappropriate use of OCPs by males?** |  |  |
| • Yes | 68 | 43.4 |
| • No | 44 | 27.7 |
| • Not sure | 46 | 28.9 |
| **What was the kind of OCPs' male customers in the last 3-6 months** |  |  |
| • Known customers | 3 | 1.9 |
| • Strangers | 43 | 27.0 |
| • Known and strangers | 28 | 17.6 |
| • Not sure | 84 | 52.8 |
| **How many males came to your pharmacy asking for OCPs in the last 3-6 months** |  |  |
| • <5 | 120 | 75.9 |
| • >5 | 38 | 24.1 |
| **How could you recognize the males who use OCPs inappropriately** |  |  |
| • From the facial impressions and body language | 43 | 27.2 |
| • They admit their needs directly | 34 | 21.5 |
| • Hard to recognize | 60 | 38.0 |
| • All the options | 21 | 13.3 |

Multivariate logistic regression results exhibited a positive significant correlation (*p*< 0.05) between the pharmacist exposure to males OCPs abuse cases and the following variables: age of the pharmacist, female gender, the length of pharmacist experience (in years), the relatively low social class distribution of the pharmacy customers, Table 4.

**Table 4.** Summary of the linear regression analysis (single and multivariant) to assess predictors associated with the exposure of participating pharmacists to OCPs use among males.

| Independent factors | Single Linear regression |  | Multivariant regression |  |
| --- | --- | --- | --- | --- |
|  | Beta | *p*-value | Beta | *p*-value |
| **Age** |  |  |  |  |
| • ≤40 years | Reference |  |  |  |
| • >40 years | 0.512 | **0.033** | 0.499 | **0.047** |
| **Gender** |  |  |  |  |
| • Female | Reference |  |  |  |
| • Male | -0.143 | **0.001** | -0.128 | **<0.001** |
| **Education** |  |  |  |  |
| • ≥ University degree | Reference |  |  |  |
| • < University degree | 0.711 | **0.127** | ----- | ----- |
| **Years of experience** |  |  |  |  |
| • ≤10 years | Reference |  |  |  |
| • >10 years | 0.349 | **0.003** | 0.325 | **<0.001** |
| **Province of the work** |  |  |  |  |
| • The capital (Amman) | Reference |  |  |  |
| • Outside the capital | -0.473 | 0.275 | ----- | ----- |
| **Social class distribution of the pharmacy customers** |  |  |  |  |
| • Low | Reference |  |  |  |
| • Moderate/high | -0.067 | **0.005** | -0.063 | **<0.001** |
| **Presence of Sport Gym around the pharmacy** |  |  |  |  |
| • Yes | Reference |  |  |  |
| • No | -0.352 | **0.052** | ----- | ----- |

**Significance ( $p < 0.05$ ) presented in bold numbers.**

### 3.4 Qualitative phases results

#### Theme 1: The most common patterns of OCPs abuse among males

Pharmacists confirmed that most of the male OCPs users were 22-35 years old. The majority of the participants highlighted that most of the males use OCPs for bodybuilding purposes, according to recommendations by their coaches at the gym. They also reported that the presence of a gym facility around the pharmacy increases the potential to observe such cases. All male customers were asking for OCPs containing estrogen (Ethinyl estradiol) and progestins (Drospirenone or Levonorgestrel). Participant 4 declared that “*After a period of 3 months, the same customers came to the pharmacy asking for Tamoxifen* (*selective estrogen receptor modulator*) *or Clomiphene citrate tablets* (*nonsteroidal, ovulatory stimulant*), *claiming that this will clear their bodies from the side effects of OCPs*”. All the participants confirmed that there is general use for hormonal products among gyms, including OCPs and testosterone.

Thirty percent of the participating pharmacists said some of the customers came to their pharmacy asking for OCPs for drug-addiction-related issues. Participant 5 stated, “*most commonly, cocaine abusers came to ask for OCPs approximately five hours before their annual urine test of illicit drugs in order to give a negative result “*. Participant 7 also mentioned that “*some customers usually asking for eye drops containing Naphazoline HCl and Chlorpheniramine Maleate to manage their eye symptoms due to addiction”*.

On the other hand, the pharmacists indicated that males use OCPs for hair growth purposes. Participant 10 said, “*As the men’s beauty centers recommend males to grind OCPs and mix them with shampoo for topical application to improve their hair growth “*. Participant 13 acknowledged “*Such practice is more common in the presence of beauty centers around the pharmacy”*. As well, fifty percent of the participants stated that beauty centers also recommend topical application of OCPs (grinded pills mixed with topical products) for acne treatment.

#### Theme 2: The role of pharmacist and pharmacy policymakers in the management of OCPs abuse among males

Most of the participants said that they could distinguish the abusers of OCPs by their facial impressions and body language/shape, as well as frequent visits to the pharmacy asking for the same products and even sometimes the customer explicitly clarify his need. Additionally, all participants agreed on the importance of the pharmacist role to fight the abuse of OCPs and came up with some recommendations such as” *Once I know that there is any misuse for OCPs, I would advise the customer to avoid this practice and mention the side effects* (*especially the risk for impotence*) *for him*,” said participant 20. In addition, they advocated the conduction of awareness campaigns, videos and distribution of pamphlets to increase knowledge and awareness about the proper use of OCPs, especially for gyms and beauty centers. Finally, the participant emphasized that pharmacists should report the abuse or misuse cases to the policymakers such as the Jordanian Pharmacist Association (JPA) and pharmacovigilance committee in the Jordan Food and drug association (JFDA) to monitor such practice.

## 4. Discussion

The abuse of body performance- or appearance-enhancing drugs by males has been increasing all over the world [16, 17], as well as in the Middle Eastern countries [17], which mostly include androgens [17, 18] and growth hormones [19]. However, the abuse of OCPs for these aforementioned purposes has never been reported in the literature. In the current study, community pharmacists were able to identify and report unanticipated patterns of OCPs use among Jordanian males, which provides an insight into the pharmacists’ role in patient education to limit medication abuse.

Half of the pharmacists participating in this study were aware of the possibility of OCPs use by males for body appearance enhancing purposes such as muscle augmenting, hair growth as well as an acne treatment. They were also knowledgeable about the side effects of OCPS in males, such as gynecomastia, mood changes, altered libido, and weight gain. These side effects were actually similar to those reported in a recent study that evaluated hormonal contraception for men [20]. Most males buying OCPs were young (25-35 year old), and this was comparable to other studies that evaluated the abuse of androgenic steroids by male athletes in Jordan which found that the mean age of the abusers was 28.1 years [21]. Tahtamouni [21] showed that in 42.9% of the abuse cases, gym coaches were responsible for recommending medication use/abuse. Similar to our study, participants also confirmed that the presence of a gym facility around the pharmacy increases the potential to observe such cases. The accessibility and affordability of OCPs make them more prone to misuse/abuse [22].

Although our findings demonstrated that around forty percent of the pharmacists refused to dispense OCPs in cases of suspected improper use, yet the rest of them would agree to do so. Unfortunately, community pharmacists in Jordan still dispensed OCPs without prescription, which is contrary to the national regulations by the JPA or JFDA [23]. The lack of control over OCPs prescribing will lead to the augmentation of the problem of improper OCPs use. Previous studies showed that these practices can be attributed to the pressure on the community pharmacists to sell the medication, due to financial considerations of the pharmacy owners, or the continuous push from the pharmaceutical companies [24]. This issue is very important at the national as well as the international level, hence, many studies have investigated the possibility of changing the status of OCPs to be an over the counter medication and expanding the scope of pharmacists’ practice beyond counseling and education to prescribing [25-28]. This suggests that there is a need for strict regulations and guidance to control any possibility of improper medication use. Still, our study showed that those pharmacists who refused to dispense emphasized that pharmacists should report the abuse or misuse cases to the policymakers such as the JPA and pharmacovigilance committee in the JFDA to monitor such practice.

Remarkably, most of the documented cases of the improper use of OCPs in this study have been rarely recognized in previous studies. Our qualitative analysis showed that OCPs were abused to mask the results of addiction to drug urine tests. It was shown that several adulterants could be added to urine in order to give a false-negative drug test, which could deter the ability to monitor illicit drug use [29]. These can include oxidizing chemicals, such as nitrite or peroxide, as well as non-oxidizing chemicals. Hajhashemi et al. (2007) conducted an in vitro and in vivo study assessing the interaction of OCPs (ethinylestradiol, levonorgestrel (LN), and both of them) at a high dose with a urine morphine diagnostic test, after reporting plenty of claims about this issue [30]. The results of that study confirmed the absence of such an interaction, which strongly suggests there is a need to stop misusing these medications. However, such practices are still available according to our study findings, which should highlight the need for further future studies to better understand this issue and devise suitable recommendations for policymakers.

A few participants reported the use of clomiphene to manage the side effects of OCPs. While this was never reported in the literature, one case report showed that a young man admitted suffering from severe depression and libido due to steroid abuse and was using clomiphene to alleviate these adverse effects [31]. Clomiphene is used in females to induce ovulation, and also has an off-label use for the treatment of hypogonadism [32, 33]. It has both estrogenic and antiestrogenic properties and initiates a series of endocrinologic events that eventually lead increase in steroidogenesis [33]. Still, the mechanism by which clomiphene can alleviate the adverse effects of OCPs is yet unclear. In this study, the reporting of OCPs abuse cases by males was correlated to many factors according to our study results, including the extent of the pharmacist’s experience. Usually, the experience in the pharmacy profession may strengthen the potential of the pharmacist to distinguish the cases of proper/abuse use of medication [34]. Moreover, the low social class distribution of pharmacy customers was one of the significantly affecting factors. The effect of socio-economic distribution on substance use (e.g. illicit drugs and alcohol) was investigated in the literature, not about OCPs, which reveals that “low social status report more environmental challenges and less psychosocial resources and that this can lead to feelings of hopelessness and a loss of coping ability” [35]. This could explain the abuse of OCPs by males to mask the urine test results of the illicit drug.

In Jordan, the policymakers and stakeholders (JFDA) emphasized that birth control pills should be handled under prescription, and till the time of writing this study it is not allowed to handle such medication as over-the-counter medications [36]. Nevertheless, a lack of surveillance and strict control opens the chance to abuse the OCPs and make them accessible to the abusers, which could ultimately lead to an increase of side effects and complications [37]. However, there is limited documentation on cases of the use and abuse cases of OCPs among males.

## 5. Strengths and limitations

This has been the first study in Jordan to addresses various patterns of use/abuse of OCPs by males, which are encountered in the community pharmacy setting. However, a number of limitations have been raised. The first limitation of this study was the survey was conducted online due to the COVID-19 pandemic that started around January of 2020 [38], along with the lockdown which was enforced by the law (till mid-June 2020). Another limitation was the use of convenient non-random sampling. Finally, this study had a small sample size (quantitative part), which might be due unfamiliarity of the pharmacists about this issue.

## 6. Conclusion

This study provided insight into unexpected uses of OCPs by males in Jordan. Community pharmacists have a crucial role in the management of OCPs use and abuse, However, restricted regulations and monitoring must be released and implemented on the community to limit such practices. These regulations should be highlighted by both policymakers and drug regulatory institutions in Jordan. In addition, there is a deep need for national educational and awareness programs for the Jordanian community about the safe and proper use of OCPs.

## Data Availability

The data will be made available by the corresponding author upon request.

## 9. Availability of data and materials

The data will be made available by the corresponding author upon request.

## References

1. WHO. Abuse (drug, alcohol, chemical, substance or psychoactive substance) 2016. Available from: https://www.who.int/substance_abuse/terminology/abuse/en/.

2. Ciccarone D. Stimulant abuse: pharmacology, cocaine, methamphetamine, treatment, attempts at pharmacotherapy. Primary Care: Clinics in Office Practice. 2011;38(1):41–58.

3. Fay JJ, Patterson D. Chapter 18 - Substance Abuse. In: Fay JJ, Patterson D, editors. Contemporary Security Management (Fourth Edition): Butterworth-Heinemann; 2018. p. 391–411.

4. Roerig JL, Steffen KJ, Mitchell JE, Zunker C. Laxative abuse. Drugs. 2010;70(12):1487–503.

5. Daniels K, Dougherty J, Jones J. Current contraceptive status among women aged 15-44: United States, 2011-2013: US Department of Health and Human Services, Centers for Disease Control and …; 2014.

6. Cooper DB, Mahdy H. Oral contraceptive pills. 2019.

7. Schindler AE. Non-contraceptive benefits of oral hormonal contraceptives. International journal of endocrinology and metabolism. 2013;11(1):41–7. Epub 2012/12/21. doi: 10.5812/ijem.4158. PubMed PMID: 23853619.

8. Kirkpatrick K. What if a man takes birth control pills? 2015 [cited 2020]. Available from: https://science.howstuffworks.com/science-vs-myth/what-if/what-if-man-takes-birth-control-pills.htm.

9. Bardaweel SK, Akour AA, Kilani M-VZJBwsh. Current knowledge, attitude, and patterns of oral contraceptives utilization among women in Jordan. 2015;15(1):117.

10. Kridli S, Newton SJINR. Jordanian married Muslim women’s intentions to use oral contraceptives. 2005;52(2):109–14.

11. Albsoul-Younes A, Wazaify M, Yousef A-M, Tahaineh LJSu, misuse. Abuse and misuse of prescription and nonprescription drugs sold in community pharmacies in Jordan. 2010;45(9):1319–29.

12. Barakat M, Al-Qudah Ra, Akour A, Al-Qudah N, Dallal Bashi YH. Unforeseen uses of oral contraceptive pills: Exploratory study in Jordanian community pharmacies. PLOS ONE. 2020;15(12):e0244373. doi: 10.1371/journal.pone.0244373.

13. Boynton PM, Greenhalgh T. Selecting, designing, and developing your questionnaire. Bmj. 2004;328(7451):1312–5.

14. Tabachnick BG, Fidell LS, Ullman JB. Using multivariate statistics: Pearson Boston, MA; 2007.

15. Bahr SJ, Hoffmann JPJT hod, society. Social scientific theories of drug use, abuse, and addiction. 2016:197–217.

16. Kanayama G, Pope HG, Jr. Illicit use of androgens and other hormones: recent advances. (1752-2978 (Electronic)).

17. Sagoe D, Pallesen S. Androgen abuse epidemiology. Current opinion in endocrinology, diabetes, and obesity. 2018;25(3):185–94. doi: 10.1097/MED.0000000000000403. PubMed PMID: 29369917.

18. Handelsman DJ. Use, Misuse, and Abuse of Androgens. In: Simoni M, Huhtaniemi IT, editors. Endocrinology of the Testis and Male Reproduction. Cham: Springer International Publishing; 2017. p. 1251–85.

19. Brennan BP, Kanayama G, Hudson JI, Pope HG, Jr. Human growth hormone abuse in male weightlifters. The American journal on addictions. 2011;20(1):9–13. doi: 10.1111/j.1521-0391.2010.00093.x. PubMed PMID: 21175915; PubMed Central PMCID: PMC3104052.

20. Thirumalai A, Ceponis J, Amory JK, Swerdloff R, Surampudi V, Liu PY, et al. Effects of 28 Days of Oral Dimethandrolone Undecanoate in Healthy Men: A Prototype Male Pill. The Journal of clinical endocrinology and metabolism. 2019;104(2):423–32. doi: 10.1210/jc.2018-01452. PubMed PMID: 30252061; PubMed Central PMCID: PMC6306388.

21. Tahtamouni LH, Mustafa NH, Alfaouri AA, Hassan IM, Abdalla MY, Yasin SR. Prevalence and risk factors for anabolic-androgenic steroid abuse among Jordanian collegiate students and athletes. European journal of public health. 2008;18(6):661–5. doi: 10.1093/eurpub/ckn062. PubMed PMID: 18603598.

22. Bardaweel SK, Akour AA, Al-Muhaissen S, AlSalamat HA, Ammar K. Oral contraceptive and breast cancer: do benefits outweigh the risks? A case -control study from Jordan. BMC Womens Health. 2019;19(1):72. doi: 10.1186/s12905-019-0770-x. PubMed PMID: 31159800; PubMed Central PMCID: PMC6547482.

23. The United States Agency for International Development (USAID). Jordan Program Profile 2015 [cited 2020 24th August]. Available from: https://www.shopsplusproject.org/sites/default/files/resources/Jordan%20Program%20Profile_final_print.pdf.

24. Mahmoud MA, Aldhaeefi M, Sheikh A, Aljadhey H. Community pharmacists perspectives about reasons behind antibiotics dispensing without prescription: a qualitative study. 2018.

25. Irwin AN, Stewart OC, Nguyen VQ, Bzowyckyj AS. Public perception of pharmacist-prescribed self-administered non-emergency hormonal contraception: An analysis of online social discourse. Res Social Adm Pharm. 2019;15(6):650–5. doi: 10.1016/j.sapharm.2018.08.003. PubMed PMID: 30143467.

26. Rafie S, Richards E, Rafie S, Landau SC, Wilkinson TA. Pharmacist Outlooks on Prescribing Hormonal Contraception Following Statewide Scope of Practice Expansion. Pharmacy (Basel). 2019;7(3). doi: 10.3390/pharmacy7030096. PubMed PMID: 31323818; PubMed Central PMCID: PMC6789671.

27. Kennedy CE, Yeh PT, Gonsalves L, Jafri H, Gaffield ME, Kiarie J, et al. Should oral contraceptive pills be available without a prescription? A systematic review of over-the-counter and pharmacy access availability. 2019;4(3):e001402.

28. McIntosh J, Rafie S, Wasik M, McBane S, Lodise NM, El-Ibiary SY, et al. Changing oral contraceptives from prescription to over-the-counter status: an opinion statement of the Women’s Health Practice and Research Network of the American College of Clinical Pharmacy. 2011;31(4):424–37.

29. Fu S. Adulterants in Urine Drug Testing. Adv Clin Chem. 2016;76:123–63. doi: 10.1016/bs.acc.2016.05.003. PubMed PMID: 27645818.

30. Hajhashemi V, Minaiyan M, Saberian-Boroojeni M. In vitro and in vivo interaction of oral contraceptive high dose (HD) with urine morphine diagnostic test. J Physiology and Pharmacology. 2007;11(1):68–75.

31. Tan RS, Vasudevan D. Use of clomiphene citrate to reverse premature andropause secondary to steroid abuse. (0015-0282 (Print)).

32. Soares AH, Horie NC, Chiang LAP, Caramelli B, Matheus MG, Campos AH, et al. Effects of clomiphene citrate on male obesity-associated hypogonadism: a randomized, double-blind, placebo-controlled study. International journal of obesity. 2018;42(5):953–63. doi: 10.1038/s41366-018-0105-2. PubMed PMID: 29777228.

33. Herzog BJ, Nguyen HMT, Soubra A, Hellstrom WJG. Clomiphene Citrate for Male Hypogonadism and Infertility: An Updated Review. Androgens: Clinical Research and Therapeutics. 2020;1(1):62–9. doi: 10.1089/andro.2020.0005.

34. Ilardo ML, Speciale AJIJoER, Health P. The Community Pharmacist: Perceived Barriers and Patient-Centered Care Communication. 2020;17(2):536.

35. Spooner C, Hetherington K. Social determinants of drug use: National Drug and Alcohol Research Centre, University of New South Wales …; 2005.

36. Jordan, Food and Drug Administration (JFDA). List of Non-prescription medications 2018 [cited 2020 24th August]. Available from: http://www.jfda.jo/Pages/viewpage.aspx?pageID=359.

37. Huang Q, Chai X, Xiao C, Cao X. A case report of oral contraceptive misuse induced cerebral venous sinus thrombosis and dural arteriovenous fistula. Medicine (Baltimore). 2019;98(33):e16440. doi: 10.1097/MD.0000000000016440. PubMed PMID: 31415348; PubMed Central PMCID: PMC6831267.

38. Lai C-C, Shih T-P, Ko W-C, Tang H-J, Hsueh P-R. Severe acute respiratory syndrome coronavirus 2 (SARS-CoV-2) and corona virus disease-2019 (COVID-19): the epidemic and the challenges. International journal of antimicrobial agents. 2020:105924.

